# Biomarker-assessed passive smoking in relation to cause-specific mortality: pooled data from 12 prospective cohort studies comprising 36 584 individuals

**DOI:** 10.1101/2020.06.04.20121111

**Authors:** Elisabeth Kvaavik, Aage Tverdal, G. David Batty

**Affiliations:** Department of Alcohol, Tobacco and Drugs, Norwegian Institute of Public Health, Oslo, Norway; Centre for Fertility and Health, Norwegian Institute of Public Health, Oslo, Norway; Department of Epidemiology and Public Health, University College London, London, UK

**Author notes:** Correspondence to: Elisabeth Kvaavik, Norwegian Institute of Public Health, Department of Alcohol, Tobacco and Drugs, PO Box 222, Skøyen, N-0213 Oslo, Norway. [,].

**Keywords:** salivary cotinine, passive smoking, mortality, cardiovascular disease, cancer

## Abstract

**Aims:** While investigators have typically quantified the health risk of passive smoking by utilising self-reported exposure, prospective studies with objective ascertainment, which are less liable to measurement error, are rare. Using data pooling, we examined the relation of a biochemical assessment of passive smoking, salivary cotinine, with mortality from a range of causes.

**Methods:** We combined data from twelve cohort studies from England and Scotland initiated between 1998 and 2008. Study members were linked to national death registries. A total of 36 584 men and women aged 16 to 85 years of age reported that they were non-smoking at baseline, provided baseline salivary cotinine, and consented to mortality record linkage.

**Results:** A mean of 8.1 years of mortality follow-up of 36 584 non-smokers (16 792 men and 19 792 women) gave rise to 2367 deaths (775 from cardiovascular disease, 780 from all cancers, and 289 from smoking-related cancers). After controlling for a range of covariates, a 10 ng/ml increase in salivary cotinine level was related to an elevated risk of total (hazard ratios; 95% confidence interval: 1.46; 1.16, 1.83), cardiovascular (1.41; 0.96, 2.09), cancer (1.49; 1.00, 2.22) and smoking-related cancer mortality (2.92; 1.77, 4.83).

**Conclusions:** Passive smoking assessed biomedically was a risk factor for a range of health outcomes known to be causally linked to active smoking.

## Introduction

Although there have been substantial secular declines in smoking prevalence in adults in the western world in recent years, more than 7 million UK adults still engage in the habit (1) and there are estimated to be over one billion smokers worldwide (2). Consequently, 13% of non-smokers report being passively exposed to tobacco smoke in home and 10% in the workplace (3).

Globally each year more than one million individuals are thought to die from passive smoking, also known as second-hand smoking or environmental tobacco smoke (2). Such estimates are based on the numerous studies that have explored the health consequences of passive smoking with the suggestion that it is linked to most diseases known to be caused by active smoking, including cardiovascular disease and selected cancers (4). In these studies, investigators have typically relied upon self-reported measures of passive smoking, raising concerns regarding reporting error, and often using a case-control design (5-8), implicating reverse causality. While more recent reports have utilised biomarkers of passive smoking in analyses of mortality or morbidity cohort data (9-11), interpretation of these findings is hampered by the small size of most studies. Accordingly, we pooled data for non-smokers across 12 cohort studies to explore the relation of salivary cotinine, a widely used indicator of passive smoking (12), with the risk of cause-specific mortality.

## Methods

The Scottish Health Survey (SHS) (13) and the Health Survey for England (HSE) (14) are independent, near-identical, cross-sectional, general population-based studies examining individuals living in households in each country. Described extensively elsewhere (15, 16), in the present analyses study members were aged 16-85 years at recruitment and were subsequently linked to a national cause-of-death registry. For the present analyses, we utilised only those surveys with salivary cotinine data (SHS 1998 and 2003; HSE 1997–2004, 2007, and 2008). Participants gave full consent and ethical approval was provided by the London Research Ethics Council.

### Assessment of active and passive smoking

Data on self-reported smoking were collected using standard enquiries (current, former and never smokers). For the cotinine assessment, a dental roll saturated with participant saliva was later analysed using a gas chromatographic method (Hewlett Packard HP5890) with a lower limit of detection of 0.1 ng/ml (levels below 0.1 ng/ml are regarded as being undetectable (17)). In the 2008 HSE this methodology was changed to a liquid chromatography tandem mass spectrometry method (18) but the two methods produce comparable results (18, 19). Salivary cotinine, a metabolite (and anagram) of nicotine, is shown to be a valid marker of tobacco smoke exposure in the previous 72 hours and to show sufficiently high specificity and sensitivity for the purposes of population-based research (20, 21).

### Assessment of confounders

Self-reported confounding factors were sex, age, survey, socio-economic status, longstanding illness, and alcohol consumption. Study member occupation was coded according to the Registrar General classification for social class (22), a six-level indicator of socio-economic status in which a lower score indicates greater prestige. Respondents reported if they suffered from a longstanding illness and their level of alcohol consumption (consumption at least 5 occasions per week was denoted as high).

### Ascertainment of cause of death

Cause of death was based on certification and coded according to the *International Classification of Diseases* (ICD, 10^th^ revision) (23). We generated outcomes for mortality from all-causes, cardiovascular disease (ICD codes I01-I99), and all cancers combined (C00-D48). Based on existing evidence (24, 25), we also denoted smoking-related cancers as C01-C16, C22, C25, C30.0, C31; C32, C34, C53, C64-C67, C68.0, C68.1, C68.8, C68.9 and C92.

### Statistical analyses

Altogether, 142 150 men and women were surveyed in 12 studies. Of these, 61 740 provided a salivary cotinine and 57 284 gave consent to use their data. After omitting self-reporting smokers (N=12 862), participants with a cotinine value at or above 15 ng/ml (self-reported non-smokers with salivary cotinine >= 15.00 ng/ml were regarded as deceivers) (N=1 971) (26, 27), and those without complete covariate data (N=5 867), 36 584 study members (19 792 women) remained. This was our analytical sample.

Having ascertained that the proportional hazards assumption had not been violated, we used Cox proportional hazards regression models (28) to compute hazard ratios (HR) with 95% confidence intervals (95% CI) to summarise the relationship between salivary cotinine level and risk of death from each mortality outcome. In these analyses, calendar time (months) was the time scale, with censoring taking place on date of death or end of mortality surveillance (February 15, 2011 for Health survey for England and December 31, 2009 for the Scottish Health Survey) - whichever came first. As there was no effect modification of the cotinine–death relation by sex or age, we combined men, women, and all ages in the analyses and adjusted for sex and age in addition to other covariates. We included survey year as fixed effects in the models. We entered two sets of covariates into the models: sex and age (comparator model); and sex, age, survey year, social class, frequency of alcohol use, and long-standing illness. The mkspline procedure in STATA produced multivariable-adjusted spline curves for any death and deaths from cardiovascular disease, total cancer, and smoking-related cancer. We carried out all analyses using Stata version 14.1 (29).

## Results

Age, sex, social status, self-reported illness and former smoking varied with cotinine level such that study members in the lowest cotinine tertile were older and there was a lower proportion of men, manual workers and former smokers relative to the higher tertiles (supplemental table). Further, higher proportions of participants in the lowest tertile reported longstanding illness than in the other groups. There was no difference in the prevalence of high alcohol intake according to cotinine categories.

The 36 584 non-smoking study participants were followed up for a mean of 8.1 years (range: 0.02 to 13.1 years) giving rise to 2367 deaths (775 deaths from cardiovascular disease, 780 from all cancers, and 289 from smoking-related cancers). In table 1 we show the relation of salivary cotinine with mortality risk. The highest level of cotinine was associated with elevated rates of death from all-causes (hazard ratio; 95% confident interval: 1.25; 1.14, 1.38) cardiovascular disease (1.33; 1.13, 1.58), and all cancers combined (1.20; 1.01, 1.42), with the strongest effect apparent for smoking-related cancers (1.57; 1.19, 2.06). There was some attenuation of risk after adjustment for multiple cofounding factors, although statistical significance at conventional levels was retained in most analyses. Mortality by continuous cotinine showed a similar pattern with elevated risk of death from any cause, cardiovascular disease, all cancers, and smoking-related cancer

**Table 1.** Hazard ratios (95% confidence interval) for the association between salivary cotinine and mortality in 36 584 non-smokers

|  | Salivary cotinine |  |  | p-value<br>for trend | Continuous salivary cotinine |  |
| --- | --- | --- | --- | --- | --- | --- |
| Salivary cotinine (ng/ml) | Tertile 1<br>(low) | Tertile 2 | Tertile 3 |  | Per 10 ng/ml<br>increase | Per 1 SD<sup>b</sup> increase |
| Range | 0-0.1 | > 0.1-0.5 | > 0.5-14.9 |  |  |  |
| Mean | 0.03 | 0.32 | 2.01 |  |  |  |
| **Total mortality** |  |  |  |  |  |  |
| **No. of deaths** | 814 | 701 | 850 |  |  |  |
| Age- and sex-adjusted | 1 (Ref) | 0.95 (0.86, 1.05) | 1.25 (1.14, 1.38) | 0.001 | 1.74 (1.40, 2.18) | 1.09 (1.05, 1.12) |
| Multiple-adjusted<sup>a</sup> | 1 | 0.91 (0.83, 1.01) | 1.18 (1.07, 1.30) | 0.002 | 1.46 (1.16, 1.83) | 1.06 (1.02, 1.09) |
| **Cardiovascular disease mortality** |  |  |  |  |  |  |
| **No. of deaths** | 264 | 227 | 284 |  |  |  |
| Age- and sex-adjusted | 1 | 0.97 (0.81, 1.16) | 1.33 (1.13, 1.58) | 0.001 | 1.78 (1.21, 2.61) | 1.09 (1.03, 1.15) |
| Multiple-adjusted<sup>a</sup> | 1 | 0.91 (0.76, 1.09) | 1.22 (1.03, 1.45) | 0.025 | 1.41 (0.96, 2.09) | 1.05 (0.99, 1.12) |
| **Cancer mortality** |  |  |  |  |  |  |
| **No. of deaths** | 275 | 216 | 288 |  |  |  |
| Age- and sex-adjusted | 1 | 0.85 (0.71, 1.01) | 1.20 (1.01, 1.42) | 0.036 | 1.73 (1.17, 2.55) | 1.09 (1.02, 1.15) |
| Multiple-adjusted<sup>a</sup> | 1 | 0.83 (0.69, 0.99) | 1.13 (0.95, 1.34) | 0.175 | 1.49 (1.00, 2.22) | 1.06 (1.00, 1.13) |
| **Smoking-related cancer mortality** |  |  |  |  |  |  |
| **No. of deaths** | 90 | 75 | 124 |  |  |  |
| Age- and sex-adjusted | 1 | 0.90 (0.66, 1.23) | 1.57 (1.19, 2.06) | 0.001 | 3.20 (1.95, 5.22) | 1.19 (1.11, 1.28) |
| Multiple-adjusted<sup>a</sup> | 1 | 0.88 (0.64, 1.19) | 1.43 (1.08, 1.89) | 0.010 | 2.92(1.77, 4.83) | 1.17 (1.09, 1.27). |

<sup>a</sup>Adjusted for sex, age, survey, social class, longstanding illness and alcohol intake. <sup>b</sup>1 SD (standard deviation): 1.5.

Figure 1 shows the spline curves for death from all-causes, cardiovascular disease, all cancers combined, and smoking-related cancers in which we illustrate thresholds of risk in relation to cotinine levels. The curves indicate that there is an increase in mortality from all causes, cancer and cardiovascular disease with increasing cotinine levels with a plateau at around 2 ng/ml such that no association seems to be present thereafter. For smoking-related cancer, this inflection is less apparent with the effect essentially being step-wise across the full cotinine range.

**Figure 1:**
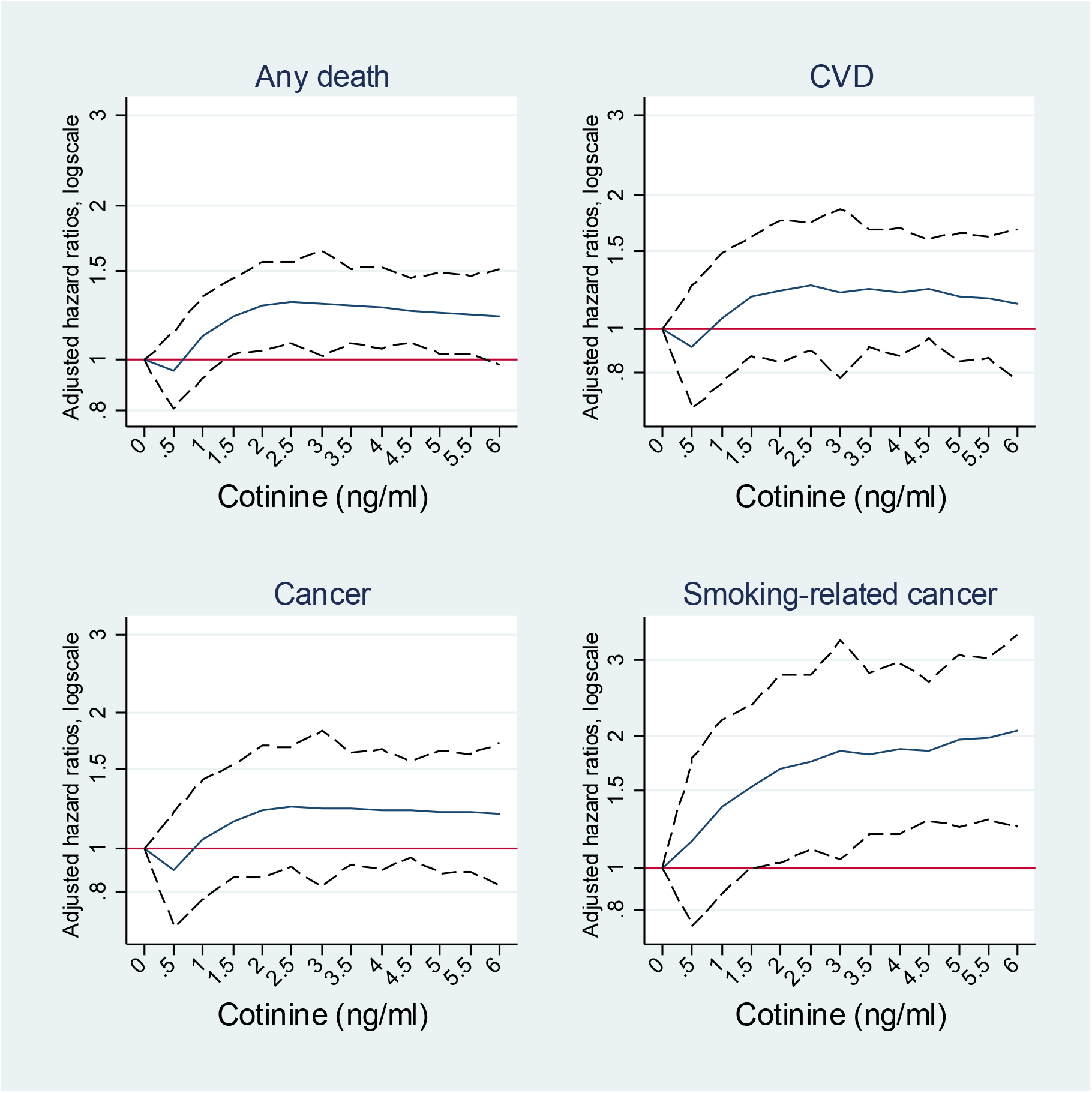
Deaths from any cause, cancer, cardiovascular disease (CVD) and smoking-related cancer by salivary cotinine level (ng/ml) Footnote: Solid line represents hazard ratio, dotted line represent 95 % confidence intervals

## Discussion

The main finding of the present analyses was that salivary cotinine, our biomarker of passive smoking, was associated with elevated rates of mortality from various causes, effects that seem to be independent of selected confounding variables. The magnitude of these relationships were, as anticipated, lower than those apparent for active smoking.

### Comparison with published studies

As discussed, few studies of mortality risk have used biomarker measurements to capture exposure to passive smoking. That different cotinine measures have been deployed – urine, saliva, blood (9, 11, 30, 31) – complicates synthesis, although correlations amongst the passive smoking indicators is high. Weak and modest associations have been reported for cotinine indices in relation to total mortality (11, 30, 32) and lung cancer (30), while associations between cotinine level and deaths from heart disease and cardiovascular disease vary (30-32). The shape of the cotinine-cardiovascular disease association in the current study suggests a threshold at low doses after which there is no distinct increase in mortality. This observation seems to accord with extant studies (11, 30-32). By contrast, the relation between cotinine and cancers ascribed to passive smoking was incremental across the cotinine continuum.

### Study strengths and limitations

A reliance on self-report of any characteristic may be problematic as interpretation may be hampered by socially desirable responses and smoking is no exception (33, 34). Our study has the advantage of having biological measurement of passive smoking, which appears to offer higher sensitivity than cotinine from urine and serum (12), and an objective health outcome, at least for fact of death. That salivary cotinine levels correlated strongly with self-reported active smoking status such that there was a marked difference between smokers and non-smokers (results not shown) gives us confidence in our results for passive smoking.

There are of course some study limitations. Passive and active smoking were captured at a single point in time and this may have resulted in some degree of misclassification of the study participants who changed habits during follow-up – during the more than decade-long period of baseline data collection (1997 – 2008), smoking prevalence, and hence passive smoking, decreased considerably (35). It is likely that this misclassification was not systematic with respect to the outcomes under study, and, as such, we have underestimated the health risks of passive smoking. Residual confounding is a perennial limitation in observational analyses and our study is no exception. Lastly, the use of cotinine, while the most commonly utilised biomarker of passive smoking, is not without its challenges. Other exposure that may influence cotinine level such as nicotine vapour from room surfaces, clothing and dust, some foods, smokeless tobacco products (snus) and nicotine replacement therapy were not captured in the present study and so cannot be taken into account. However, we did exclude study members because their cotinine levels were too high for them to be realistically classified as non-smokers. It is possible that at least some of these people had used nicotine products other than cigarettes.

In conclusion, our study supports an association between objectively ascertained second hand smoking and mortality from any death, cardiovascular disease and smoking-related cancers. The apparent threshold effects for some of these relationships require further exploration.

## Data Availability

Data can be downloaded from the UK data archive (www.data-archive.ac.uk)

## Declaration of competing interests

None

---

**Supplementary table.** Baseline characteristics by salivary cotinine level in 36 584 non-smokers

|  |  | **Salivary cotinine (ng/ml) tertiles** |  |  |  |
| --- | --- | --- | --- | --- | --- |
|  | All (0-14.9<br>ng/ml) | Low<br>(0-0.1) | Middle<br>(> 0.1-0.5) | High<br>(> 0.5-14.9) | P-value for<br>difference<br>across tertiles |
| N | 36 584 | 13320 | 11471 | 11793 |  |
| *Mean* |  |  |  |  |  |
| Age (years) |  | 51.9 | 48.9 | 46.0 | < 0.001 |
| Cotinine (ng/ml) |  | 0.03 | 0.32 | 2.01 | < 0.001 |
| *Percentages* |  |  |  |  |  |
| Males |  | 41.7 | 44.9 | 51.6 | < 0.001 |
| Manual social<br>class |  | 30.7 | 36.0 | 46.5 | < 0.001 |
| Longstanding<br>illness |  | 46.4 | 44.7 | 43.8 | < 0.001 |
| High alcohol<br>intake |  | 18.3 | 17.3 | 18.5 | 0.06 |
| Formers smokers |  | 30.9 | 31.2 | 34.0 | < 0.001 |

## Notes

### Competing Interest Statement

The authors have declared no competing interest.

### Clinical Trial

Not applicable

### Funding Statement

No direct funding

### Author Declarations

Participants gave full consent and ethical approval was provided by the London Research Ethics Council. The present study is based on analyses of anonymized data and therefore it did not require any further approvals or permissions.

